# Hepatitis E virus seroprevalence and associated risk factors in high-risk groups: A cross-sectional study from Turkey

**DOI:** 10.1101/2021.01.25.21250429

**Authors:** H. Yasemin Balaban, Abdullah Tarık Aslan, Fatma Nur Akdoğan Kittana, Alpaslan Alp, Osman Dağ, Şefika Nur Ayar, Cavanşir Vahabov, Cem Şimşek, Tolga Yıldırım, Hakan Göker, Koray Ergünay, Yunus Erdem, Yahya Büyükaşık, Halis Şimşek

## Abstract

**Background:** The renal transplant recipients (RT), allogeneic hematopoietic stem cell transplant recipients (allo-HSCT), patients with acute hepatitis (AH), and chronic hepatitis C patients (CHC) are at risk of hepatitis E virus (HEV) infection. However, seroepidemiology, risk factors to HEV exposure, and the prevalence of HEV viremia has not yet been investigated among these patients in Turkey.

**Materials&Methods:** In this cross-sectional study, 292 consecutive serum samples were tested for HEV immunoglobulin IgG/IgM and HEV RNA using commercial ELISA and in-house nested PCR with Sanger sequencing, respectively. Sociodemographic, clinical, laboratory data, and risk factors were collected using a questionnaire and hospital database. Multiple logistic regression analysis was employed to identify independent predictors for anti-HEV seropositivity.

**Results:** Among all patients (n=292) tested for HEV RNA reactivity, only 2 patients (one RT recipient and one patient with AH) were identified as having HEV3 viremia. HEV viremia rate was 0.6% in whole group. These patients had shown no signs of chronic HEV infection for 6 months and was found to spontaneously seroconverted 6 months after enrollment. Anti-HEV IgG was positive in 29 patients yielding an HEV seroprevalence of 9.9%. Older age (aOR:1.03, 95% CI, 1.00-1.06; *p*:0.022) and eating undercooked meat (aOR:3.11, 95% CI, 1.08-8.92; *p*:0.034) were independent risk factors to anti-HEV seropositivity in all patients. Similarly, multiple logistic regression analysis demonstrated that age (aOR:1.03, 95% CI, 0.99-1.07, *p*:0.058) and eating undercooked meat (aOR:5.77, 95% CI, 1.49-22.25, *p*:0.011) were independent risk factors for anti-HEV IgG positivity in the non-immunosuppressive subgroup consisting of AH and CHC patients.

**Conclusion:** The HEV seroprevalence rate was high (9.9%), despite low viremia rate (0.6%) in high-risk patients. The emergence of HEV3 might indicate a serious problem for these patients. Future investigations are needed to elucidate foodborne transmission routes of HEV in Turkey.

## Introduction

Hepatitis E virus (HEV) is a small non-enveloped virus with a single-stranded positive sense ribonucleic acid (RNA) genome (1,2). HEV is the most frequent cause of acute viral hepatitis all over the world and classified in the genus *Hepevirus* and the family *Hepeviridae* (3-5). Among the eight distinct HEV genotypes, HEV1, HEV2, HEV3, HEV4, and recently reported HEV7 are mainly responsible for infection of humans (6). Contaminated water supply is the main source of HEV1 and HEV2 transmission to humans and transmission from animals to humans for HEV1 and HEV2 has not yet been reported (7). HEV3 and HEV4 infections develop mainly through zoonotic transmission from consumption of contaminated foodstuff (mostly raw or undercooked meat) and less frequently from direct contact with infected animals (7). Their main reservoir is the pig, but they have also been shown in wild boars, rabbits, goats, sheep, deer, horses, cats, and dogs (8-11). Albeit less common than zoonotic and waterborne transmissions, other possible transmission route for HEV is transfusions of infected blood or blood products.

HEV infections have various clinical presentations including acute and self-limiting hepatitis, acute-on-chronic liver disease, chronic hepatitis, cirrhosis, and liver failure. Additionally, HEV infection is associated with extrahepatic involvement affecting wide range of organ systems, although the causal relationship for many of them still needs to be proven (7). Infections with HEV3 and HEV4 are usually associated with silent seroconversion and mostly seen as autochthonous (locally acquired) zoonotic infections in developed countries. However, immunocompromised patients including solid organ transplant recipients, allogeneic hematopoietic stem cell transplant recipients, and patients with HIV infections have been shown to develop chronic infections with the risk of rapid progression to cirrhosis within 2-5 years (12). The major risk groups for HEV infection and associated adverse outcomes are pregnant women, infants, older people, immunocompromised individuals, patients with underlying chronic liver diseases, and workers that come into close contact with HEV-infected animals.

Several seroprevalence studies from Europe were regional and demonstrated great heterogeneity in methodology and results (13). There is also a risk of underestimation in these studies because of variations in performance of commercially available ELISA assays (14). Nevertheless, a survey conducted in 30 European countries reported a remarkable increase in the number of cases from 514 per year in 2005 to 5617 per year in 2015 (15). However, little is known about HEV seroprevalence and risk factors contributing HEV infection in Turkey. Turkey has been reported as an HEV1 and HEV2 endemic country based upon outdated data, although current epidemiologic data are lacking.

In this cross-sectional study, we aimed to determine anti-HEV IgG seroprevalence and the rate of HEV RNA positivity as well as independent risk factors for anti-HEV IgG seropositivity in special risk groups, namely renal transplant recipients (RT), allogeneic hematopoietic stem cell transplant recipients (allo-HSCT), patients with acute hepatitis (AH), and chronic hepatitis C patients (CHC).

## Material and Methods

This cross-sectional study was conducted in a 1000-bed tertiary care academic hospital (Hacettepe University Faculty of Medicine Hospital). Through the one-year period starting from 01.10.2018, adult patients (aged ≥18 years) with a history of RT, allo-HSCT, AH, and CHC were included in the study. Except one patient allocated in allo-HSCT group withdrew her informed consent after initial enrollment, there has not been any drop-out during the study period. A total of 157 patients with RT, 46 patients with allo-HSCT, 19 patients with AH, and 70 CHC patients were evaluated in the final analysis. Data pertaining with demographics, underlying disorders, family history of liver diseases, laboratory test results, presence of hepatomegaly and splenomegaly according to ultrasonographic examination, type of profession, educational level, household income per month, farm animal ownership, type of residency (rural/urban), eating undercooked meat, consumption of drinking raw milk, transfusion of blood or blood products, immunosuppressive drug use, previous history of hemodialysis and surgical intervention, and past ribavirin exposure were collected through both face to face interviews and from the hospital database.

Sera obtained from 292 participants were stored at -80°C. Screening for anti-HEV IgM and IgG antibodies was carried out by commercial ELISA tests (HEV semi-quantitative IgM and quantitative IgG, Euroimmun, Germany), performed according to the manufacturer instructions. For nucleic acid purification and subsequent cDNA synthesis, commercial assays (Gene MATRIX viral RNA/DNA purification and EurX smART first strand cDNA synthesis, EurX, Poland) were used as directed by the manufacturer. Detection of HEV genome was carried out by partial amplification of the ORF2 region by a nested polymerase chain reaction (PCR) assay, performed as described previously (16). PCR products were cleaned up using High Pure PCR product purification kit (Roche Diagnostics, Germany) and sequenced in an ABI Prism 310 Genetic Analyzer (Applied Biosystems, USA.). Obtained sequences were handled using Geneious software v11.1.5 (Biomatters Ltd., New Zealand). Nucleotide similarity searches were carried out in the National Center for Biotechnology Information website, using BLASTn and BLASTn optimized for highly similar sequences (MEGABLAST) algorithms (17). Nucleotide and putative amino acid alignments and pairwise sequence comparisons were performed using CLUSTAL W (18). Evolutionary history was inferred via the maximum-likelihood method, based on the estimated optimal substitution model individually for each alignment according to the Bayesian information criterion and conducted using MEGAX (19). Individuals with detectable viremia were followed up for up to 6 months for clinical symptoms, laboratory and HEV tests (RNA and IgG).

## Ethics

The ethical approval for this study was obtained from Hacettepe University local ethics committee (study approval identification code: GO 18/186). The study was conducted accordance with the Declaration of Helsinki and national and institutional standards. Patients providing informed consent were enrolled in this study. No fee was paid to the participants. All data were anonymously collected on a secured dataset by the primary investigators without any identifying data.

## Statistical Analysis

Descriptive statistics were reported to demonstrate the baseline characteristics of anti-HEV seropositive and seronegative groups. For continuous variables, mean±standard deviation values were reported when the certain assumptions such as normality and variance homogeneity are satisfied; otherwise, median (minimum-maximum) values were presented. The normality and variance homogeneity assumptions were assessed by applying Shapiro-Wilk test and Levene’s test, respectively. Independent t-test was used for comparing two groups when these assumptions were satisfied. Otherwise, Mann-Whitney U test was applied. Frequency (%) was stated for categorical variables. While comparing the two distinct groups in terms of categorical variables, Pearson chi-square test was used if the number of observations is large enough in each cell of cross table; otherwise, Fisher’s exact test was conducted. Odds ratios were utilized for comparing two groups with respect to investigated outcomes. Multiple logistic regression analysis was run to identify factors being associated with HEV seropositivity. Variables with a *P* value of ≤ 0.1 in univariate analysis were incorporated into multiple logistic regression model. Hoshmer-Lemeshow test was used to assess the goodness of fit of the model. Youden’s J statistic was utilized to select the optimal cut-point used in the multiple logistic regression analysis. Multicollinearity analysis and correlation analysis were used to clear possible collinearity and correlation issues among variables put into multiple logistic regression model. A two-sided p value ≤ 0.05 was considered significant. All statistical analyses were run using Statistical Package for Social Sciences (SPSS Inc, Chicago, IL, USA) 23.0 version for Windows.

## Results

Baseline characteristics of patients are shown in Table 1. Among the 292 patients, 29 patients were anti-HEV IgG positive, yielding an HEV seroprevalence of 9.9%. The HEV IgG positivity rates were 5.4% and 20.2% in immunocompromised and non-immunocompromised groups, respectively (p <0.001). None of the anti-HEV IgG positive patients was positive for anti-HEV IgM. Among all the risk factors analyzed, age and eating undercooked meat were the only risk factors maintaining statistical significance in multiple logistic regression analysis (Table 2). For all patients, the source of drinking water was public water supply system, and the household sewage system was connected to public sewage system. Also, none of the patients stated a habit of consuming pork. Therefore, the impact of these possible risk factors on HEV seroprevalence could not be evaluated. The risk factor analyzes were also carried out for non-immunocompromised patients including AH and CHC subgroups. In univariate analysis, age, low educational level, lower GGT level (IU/ml), and eating undercooked meat seemed to be associated with anti-HEV IgG seropositivity (table 3). Consistent with the analysis encompassing all patients, multiple logistic regression analysis revealed that older age and eating undercooked meat were independent risk factors for anti-HEV IgG positivity among non-immunocompromised patients (Table 3). A similar subgroup analysis for immunosuppressive patients including RT and allo-HSCT groups could not be performed because of very low number of anti-HEV IgG positive patients in these groups.

**Table 1.** Baseline features of all patients according to Anti-HEV IGG status

| Characteristics | All patient groups (n=292) | Anti-HEV IGG positive (n=29) | Anti-HEV IGG negative (n=263) | p-value* |
| --- | --- | --- | --- | --- |
| Age (years) | 47.0 (18-89) | 59.0 (18-81) | 46.0 (18-89) | **<0.001<sup>c</sup>** |
| Male Sex, n (%) | 143 (48.9) | 13 (44.8) | 130 (49.4) | 0.78<sup>a</sup> |
| Educational level, n (%) |  |  |  | **< 0.001<sup>b</sup>** |
| • Illiterate | 15 (5.1) | 5 (17.2) | 10 (3.8) |  |
| • Preliminary school | 134 (45.8) | 19 (65.5) | 115 (43.7) |  |
| • High school | 68 (23.2) | 2 (6.9) | 66 (25.1) |  |
| • University/college | 68 (25.6) | 3 (10.3) | 72 (27.4) |  |
| Rural residence, n (%) | 50 (17.2) | 8 (27.6) | 42 (16.0) | 0.18<sup>a</sup> |
| Animal ownership, n (%) | 67 (22.9) | 10 (34.5) | 57 (21.7) | 0.18<sup>a</sup> |
| Ownership of farm animals, n (%) | 40 (13.6) | 8 (27.6) | 32 (12.2) | 0.06<sup>a</sup> |
| Eat undercooked meat, n (%) | 31 (10.6) | 7 (24.1) | 24 (9.1) | 0.03<sup>a</sup> |
| Drink raw milks, n (%) | 87 (29.7) | 11 (37.9) | 76 (28.9) | 0.42<sup>a</sup> |
| Alcohol consumption, n (%) | 25 (8.5) | 1 (3.4) | 24 (9.1) | 0.48<sup>b</sup> |
| Family history of liver diseases, n (%) | 9 (3.0) | 2 (6.9) | 7 (2.7) | 0.22<sup>b</sup> |
| Household income, n (%) |  |  |  | 0.38<sup>b</sup> |
| • <2000 tl | 167 (57.1) | 20 (69.0) | 147 (55.9) |  |
| • 2000-10.000 tl | 115 (39.3) | 8 (27.6) | 107 (40.7) |  |
| • >10.000 tl | 10 (3.4) | 1 (3.4) | 9 (3.4) |  |
| Primary Diseases, n (%) |  |  |  | **0.001<sup>b</sup>** |
| • Renal transplantation, n (%) | 157 (53.7) | 6 (20.7) | 151 (57.4) |  |
| • Allogeneic hematopoietic transplantation, n | 46 (15.7) | 5 (17.2) | 41 (15.6) |  |
| (%) |  |  |  |  |
| • Acute hepatitis, n (%) | 19 (6.5) | 3 (10.3) | 16 (6.1) |  |
| • Chronic hepatitis C, n (%) | 70 (23.9) | 15 (51.7) | 55 (20.9) |  |
| History of blood transfusion, n (%) | 80 (27.3) | 7 (24.1) | 73 (27.8) | 0.84<sup>a</sup> |
| Prior surgical intervention, n (%) | 220 (75.3) | 14 (48.3) | 206 (78.3) | **0.001<sup>a</sup>** |
| Previous history of hemodialysis, n (%) | 110 (37.6) | 5 (17.2) | 105 (39.9) | **0.016<sup>b</sup>** |
| Hepatomegaly, n (%) | 59 (20.2) | 5 (17.2) | 54 (20.5) | 0.81<sup>b</sup> |
| Splenomegaly, n (%) | 41 (14.0) | 3 (10.3) | 38 (14.4) | 0.77<sup>b</sup> |
| Comorbidities |  |  |  |  |
| • Previous history of acute hepatitis, n (%) | 64 (21.9) | 9 (31.0) | 55 (20.9) | 0.31<sup>a</sup> |
| • Cirrhosis, n (%) | 19 (6.5) | 4 (13.8) | 15 (5.7) | 0.10<sup>b</sup> |
| • Diabetes Mellitus, n (%) | 49 (16.7) | 4 (13.8) | 45 (17.1) | 0.79<sup>b</sup> |
| • Chronic Kidney Diseases, n (%) | 165 (56.5) | 8 (27.6) | 157 (59.7) | **0.002<sup>a</sup>** |
| • Coronary Arterial Diseases, n (%) | 24 (8.2) | 3 (10.3) | 21 (8.0) | 0.71<sup>b</sup> |
| • Respiratory Diseases, n (%) | 14 (4.7) | 2 (6.9) | 12 (4.6) | 0.63<sup>b</sup> |
| • Hematological Malignancy, n (%) | 47 (16.0) | 6 (20.7) | 41 (15.6) | 0.65<sup>a</sup> |
| • Chronic hepatitis B, n (%) | 16 (5.4) | 2 (6.9) | 14 (5.3) | 0.66<sup>b</sup> |
| Immunosuppressive Treatment |  |  |  |  |
| • Active immunosuppressive therapy use, n (%) | 172 (58.9) | 8 (27.6) | 164 (62.4) | **0.001<sup>a</sup>** |
| • Past use of immunosuppressive therapy, n (%) | 177 (60.6) | 9 (31.0) | 168 (63.9) | **0.001<sup>a</sup>** |
| • Active steroid use, n (%) | 155 (53.0) | 7 (24.1) | 148 (56.3) | **0.002<sup>a</sup>** |
| • Active mycophenolate mofetil use, n (%) | 117 (40.0) | 5 (17.2) | 112 (42.6) | **0.009<sup>b</sup>** |
| • Active tacrolimus use, n (%) | 110 (37.6) | 6 (20.7) | 104 (39.5) | 0.07<sup>a</sup> |
| • Active cyclosporine use, n (%) | 42 (14.3) | 0 (0.0) | 42 (16.0) | **0.012<sup>b</sup>** |
| • Past ribavirin exposure, n (%) | 43 (14.7) | 12 (41.4) | 31 (11.8) | **< 0.001<sup>a</sup>** |
| Tacrolimus dose (tablet/day) | 0 (0-8) | 0 (0-5) | 0 (0-8) | 0.21<sup>c</sup> |
| Duration of tacrolimus use (years) | 0 (0-16) | 0 (0-13) | 0 (0-16) | 0.09<sup>c</sup> |
| Mycophenolate mofetil dose (tablet/day) | 0 (0-4) | 0 (0-4) | 0 (0-4) | **0.009<sup>c</sup>** |
| Duration of mycophenolate mofetil use (years) | 0 (0-16) | 0 (0-5) | 0 (0-16) | **0.001<sup>c</sup>** |
| Cyclosporine A dose (tablet/day) | 0 (0-5) | 0 (0-5) | 0 (0-5) | 0.20<sup>c</sup> |
| Duration of Cyclosporine A use (years) | 0 (0-5) | 0 (0-1) | 0 (0-5) | 0.08<sup>c</sup> |
| Azathioprine dose (tablet/day) | 0 (0-2) | 0 (0-2) | 0 (0-2) | 0.52<sup>c</sup> |
| Duration of azathioprine use (years) | 0 (0-25) | 0 (0-13) | 0 (0-25) | 0.51<sup>c</sup> |
| Steroid dose (tablet/day) | 1 (0-8) | 1 (0-8) | 0 (0-2) | **0.003<sup>c</sup>** |
| Duration of steroid use (years) | 2 (0-29) | 0 (0-13) | 2 (0-29) | **0.001<sup>c</sup>** |
| Time after transplantation (years) | 5 (1-33) | 4 (1-33) | 5 (1-27) | 0.89<sup>c</sup> |
| Number of transplantations | 1 (0-3) | 0 (0-1) | 1 (0-3) | **< 0.001<sup>c</sup>** |
| Age at transplantation (years) | 36 (4-66) | 42 (6-66) | 35 (4-64) | 0.21<sup>c</sup> |
| Number of rejections | 0 (0-2) | 0 (0-1) | 0 (0-2) | 0.16<sup>c</sup> |
| Laboratory values |  |  |  |  |
| • Hemoglobin (g/l) | 12.8±2.2 | 12.9±2.1 | 12.8±2.2 | 0.77<sup>d</sup> |
| • Platelet (10<sup>3</sup>/mm<sup>3</sup>) | 218549±97529 | 212620±85928 | 219203±98850 | 0.82<sup>d</sup> |
| • ALT (U/l) | 18 (4-3736) | 18 (4-1201) | 18 (4-3736) | 0.38<sup>c</sup> |
| • AST (U/l) | 20 (4-8714) | 24 (9-494) | 20 (4-8714) | 0.16<sup>c</sup> |
| • ALP (U/l) | 85 (27-750) | 92 (47-690) | 82 (27-750) | 0.07<sup>c</sup> |
| • GGT (U/l) | 29 (7-1450) | 21 (10-1450) | 30 (7-644) | 0.37<sup>c</sup> |
| • Total bilirubin (mg/dl) | 0.69 (0.1-23.2) | 0.69 (0.31-3.9) | 0.69 (0.1-23.2) | 0.63<sup>c</sup> |
| • Albumin (g/dl) | 4.01±0.53 | 3.87±0.53 | 4.02±0.53 | 0.12<sup>c</sup> |
| • INR | 1 (0.8-3.9) | 1 (0.8-2.3) | 1 (0.8-3.9) | 0.16<sup>c</sup> |
| • Creatinine (mg/dl) | 0.9 (0.3-12) | 0.8 (0.4-7.6) | 0.9 (0.3-12) | 0.02<sup>c</sup> |

Results are reported as mean±SD for the continuous variables satisfying the normality assumption and median (min-max) for the continuous variables not satisfying the normality assumption. Categorical variables are summarized with frequency (%) of patients
ALT, alanine aminotransferase; AST: aspartate aminotransferase, ALP, alkaline phosphatase; GGT: gamma glutamyl transferase; INR, international normalized ratio, mg, milligram; g, gram; dl, deciliter; U, unite; n, sample size
a. Pearson $\chi^2$ test, b. Fisher exact test, c. Mann-Whitney U test, d. Independent sample t test
\*p value is for the difference between anti-HEV IgG positive and negative groups

**Table 2.** Risk factors for anti-HEV IgG positivity among all patients

| Variables | No of patients |  | p-value | Multivariate analysis |  |
| --- | --- | --- | --- | --- | --- |
|  | Anti-HEV IgG+<br>(n=29) | Anti-HEV IgG-<br>(n=263) |  | aOR (95% CI) | p-value |
| Median (IQR) Age (years) | 59.0 (18-81) | 46.0 (18-89) | <0.001<sup>d</sup> | 1.03 (1.00-1.06) | **.022<sup>b</sup>** |
| Educational level, n (%) |  |  | <0.001<sup>c</sup> | 0.59 (0.33-1.07) | .086<sup>b</sup> |
| • Illiterate | 5 (17.2) | 10 (3.8) |  |  |  |
| • Preliminary school | 19 (65.5) | 115 (43.7) |  |  |  |
| • High school | 2 (6.9) | 66 (25.1) |  |  |  |
| • University/college | 3 (10.3) | 72 (27.4) |  |  |  |
| Eat undercooked meat, n (%) | 7 (24.1) | 24 (9.1) | .03<sup>a</sup> | 3.11 (1.08-8.92) | **.034<sup>b</sup>** |
| Ownership of farm animals, n (%) | 8 (27.6) | 32 (12.2) | .06<sup>a</sup> | 1.06 (0.35-3.15) | .911<sup>b</sup> |
| Previous history of hemodialysis, n (%) | 5 (17.2) | 105 (39.9) | .016<sup>c</sup> | ..... | ..... |
| Prior surgical intervention, n (%) | 14 (48.3) | 206 (78.3) | .001<sup>a</sup> | ..... | ..... |
| Active immunosuppressive therapy use, n (%) | 8 (27.6) | 164 (62.4) | .001<sup>a</sup> | 1.21 (0.26-5.63) | .803<sup>b</sup> |
| Past use of immunosuppressive therapy, n (%) | 9 (31.0) | 168 (63.9) | .001<sup>a</sup> | ..... | ..... |
| Active tacrolimus use, n (%) | 6 (20.7) | 104 (39.5) | .07<sup>a</sup> | ..... | ..... |
| Active mycophenolate mofetil use, n (%) | 5 (17.2) | 112 (42.6) | .009<sup>c</sup> | ..... | ..... |
| Active steroid use, n (%) | 7 (24.1) | 148 (56.3) | .002<sup>a</sup> | ..... | ..... |
| Chronic Kidney Diseases, n (%) | 8 (27.6) | 157 (59.7) | .002<sup>a</sup> | ..... | ..... |
| Primary Diseases, n (%) |  |  | .001<sup>c</sup> |  |  |
| • Renal transplantation, n (%) | 6 (20.7) | 151 (57.4) |  | 3.61 (0.96-13.52) | .056<sup>b</sup> |
| • Allogeneic hematopoietic transplantation, n(%) | 5 (17.2) | 41 (15.6) |  | 2.11 (0.37-11.82) | .396<sup>b</sup> |
| • Acute hepatitis, n(%) | 3 (10.3) | 16 (6.1) |  | 2.61 (0.78-8.76) | .119<sup>b</sup> |
| • Chronic hepatitis C, n(%) | 15 (51.7) | 55 (20.9) |  | ref |  |
| Past ribavirin exposure, n (%) | 12 (41.4) | 31 (11.8) | <0.001<sup>a</sup> | ..... | ..... |
| ALP (U/l) | 21 (10-1450) | 82 (27-750) | .07<sup>d</sup> | 1.00 (0.99-1.00) | .945<sup>b</sup> |
| Creatinine (mg/dl) | 0.8 (0.4-7.6) | 0.9 (0.3-12) | .02<sup>d</sup> | ..... | ..... |
| Number of | 0 (0-1) | 1 (0-3) | <0.001<sup>d</sup> | ..... | ..... |
| Transplantations |  |  |  |  |  |
| Duration of steroid use, (years) | 0 (0-13) | 2 (0-29) | .001<sup>d</sup> | ..... | ..... |
| Steroid dose, (tablet/day) | 1 (0-8) | 0 (0-2) | .003<sup>d</sup> | ..... | ..... |
| Mycophenolate mofetil dose, (tablet/day) | 0 (0-4) | 0 (0-4) | .009<sup>d</sup> | ..... | ..... |
| Duration of mycophenolate mofetil use, (years) | 0 (0-5) | 0 (0-16) | .001<sup>d</sup> | ..... | ..... |
| Duration of tacrolimus use, (years) | 0 (0-13) | 0 (0-16) | .09<sup>d</sup> | ..... | ..... |
| Duration of Cyclosporine A use, (years) | 0 (0-1) | 0 (0-5) | .08<sup>d</sup> | 0.68 (0.36-1.31) | .259<sup>b</sup> |
| Abbreviations: ALP, alkaline phosphatase; IQR, interquartile range; HEV, Hepatitis E virus; IgG, immunoglobulin G; mg, milligram; dl, deciliter; U, unite; OR, odds ratio; aOR, adjusted odds ratio; CI, confidence interval; n, sample size |  |  |  |  |  |
| <sup>a</sup>P Value was obtained by Pearson chi-square test, <sup>b</sup>P Value was obtained by multivariate logistic regression analysis, <sup>c</sup>P Value was obtained by Fisher's exact test, <sup>d</sup>P value was obtained by Mann-Whitney U test |  |  |  |  |  |

Among the 292 patients, two were initially found to be HEV RNA positive. Therefore, the HEV RNA viremia rate was 0.6%. Of these two patients, one had been treated with renal transplantation and other patient had acute hepatitis. HEV RNA prevalence rate was found 0.6% in the RT group as similar with that of all patients. In patients with detectable HEV RNA, partial ORF2 sequences of 407 nucleotides were obtained by amplicon sequencing. The sequences corresponded to the 778-1184. positions on the ORF 2 (isolate HRC-HE104, GenBank accession: AB630970.1) and demonstrated 99.7% similarity with identical putative amino acid sequence. In the maximum likelihood analysis, they formed a separate cluster within genotype 3 viruses, supported by high bootstrap values (Figure 1). In both patients with initial HEV viremia, clearance of HEV viremia and anti-HEV seroconversion were detected in the follow-up samples taken at 6 months after enrollment. We concluded that the viremia by HEV genotype 3 in both patients was due to acute infections, without progressing chronic HEV infection or hepatitis.

**Figure 1.**
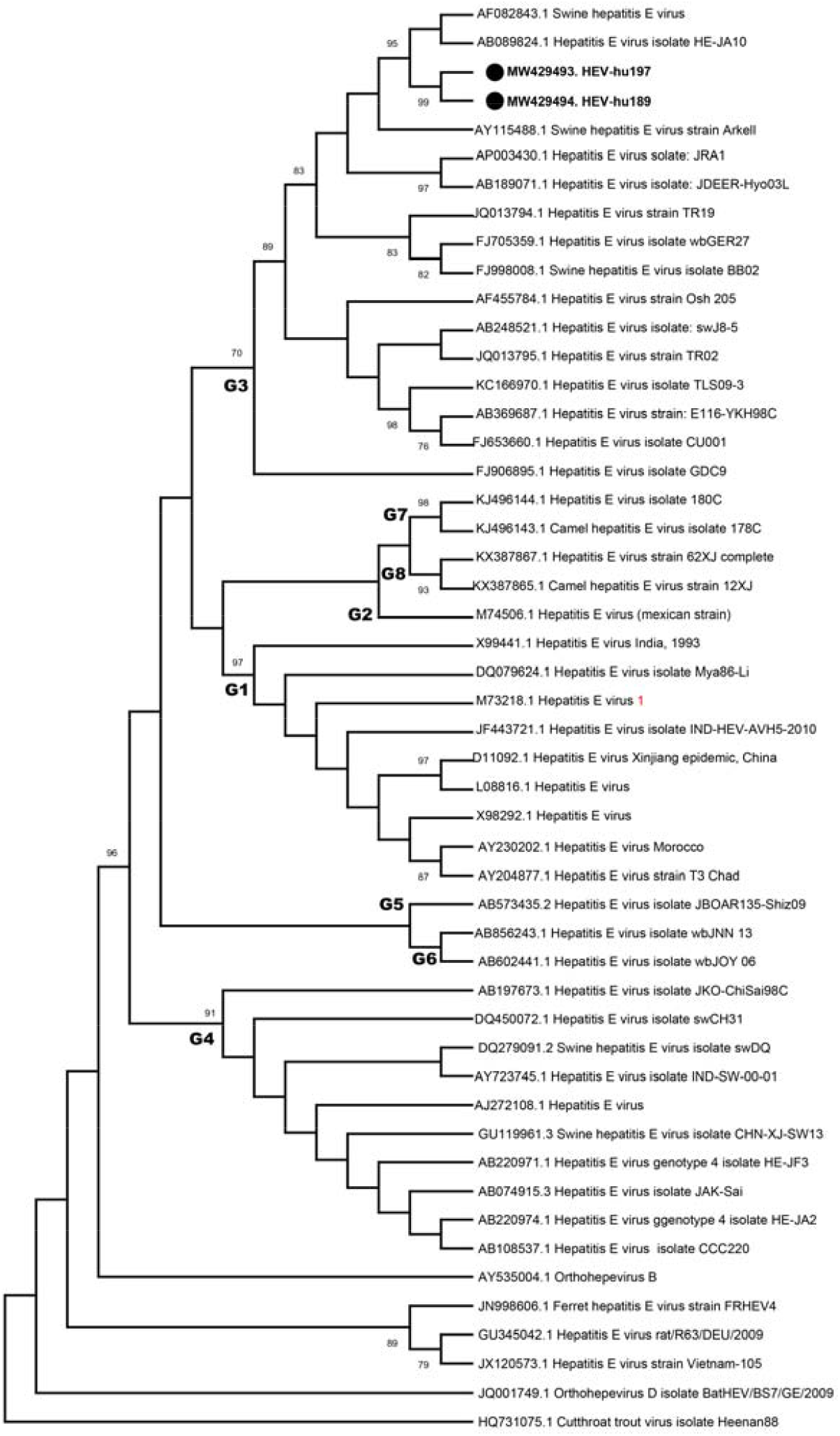
The maximum likelihood analysis of the HEV partial ORF2 sequences (407 nucleotides). The tree is constructed using the General Time Reversible (GTR) model, with a discreet Gamma distribution (+G) for 500 replications. Viruses are indicated by GenBank accession number, name and strain identifier, if available. The sequences characterized in this study are marked and indicated with GenBank accession number. Major HEV genotypes are indicated on the tree. Bootstrap values higher than 70 are provided. Cutthroat trout virus isolate Heenan88 is included as an outgroup.

### Clinical features of HEV RNA positive patients

A 39-year-old male (patient 1) has a university degree and been working as a bank clerk. He suffered from focal segmental glomerulosclerosis as a primary renal disease and underlying cause of renal insufficiency. Before renal transplantation, he had been undergone hemodialysis for 8 months. He has been treated with mycophenolate mofetil, prednisolone, and tacrolimus for 8 years due to renal transplantation. He did not have any farm animal exposure, habit of eating undercooked meat and drinking raw milk, previous acute hepatitis history, and organ rejection. He had a prior surgical intervention for renal transplantation and a history of alcohol consumption as one beer or 2 double arrack per month. In laboratory analysis, hepatic enzymes were normal (ALT:29 IU/ml, ALP:65 IU/ml, GGT:27IU/ml). Total bilirubin, albumin, creatinine, and INR values were 0.4 mg/dl, 4.6 g/dl, 1.3 mg/dl, and 0.9, respectively. He did not have immunity against HBV and the serologies of other viral hepatitis were negative. Both anti-HEV IgG and anti-HEV IgM were also negative, initially. The result of genotype analysis turned out genotype 3. He had not developed any manifestation that can be consistent with hepatic and/or extrahepatic involvement of HEV during 6 months follow-up period. The clearance of HEV viremia and anti-HEV IgG seroconversion were detected at the visit executed 6 months after diagnosis. The patient did not specify any signs or symptoms that can be associated with HEV infection and laboratory analysis was unremarkable at this visit.

A 36-year-old male (patient2) has underlying cholelithiasis and depression. He had complaints of right upper quadrant pain, nausea, and vomiting. He had jaundice and hepatomegaly in physical examination. His medical history unfolded the recent consumption of mighty pomegranate (Punica granatum). In abdomen ultrasound, CT (Computerized Tomography), and MRCP (Magnetic Resonance Cholangiopancreatography) examinations, multiple bile stones up to 2 cm in diameter, mild dilatation in intrahepatic biliary ducts, hepatomegaly, and suspected signs of cholangitis were detected. In laboratory analysis, hepatic enzymes were very high (ALT:903, ALP: 638, GGT:593). The synthesis function of liver was normal (Albumin:4.6 g/dl, INR:1). There was a mixed type of hyperbilirubinemia (total bilirubin: 7.3 mg/dl, direct bilirubin: 3.3 mg/dl). He had an immunity against HBV gained by previous exposure. Other viral hepatitis serologies were negative. In serological analysis, both anti-HEV IgG and anti-HEV IgM were negative, though genotype analysis revealed HEV genotype 3 viremia. This patient was admitted to the hospital and treated with biliary stone extraction with ERCP (Endoscopic retrograde cholangiopancreatography) and antibiotic therapy. After 7 days of treatment, all the complaints of the patient improved significantly. Similarly, the significant decline in values of hepatic enzymes and bilirubin levels at 1 week after presentation were consistent with acute resolving hepatitis. At the visit undertaken 6 months after initial presentation, both seroconversion of anti-HEV IgG and clearance of HEV viremia were identified.

## Discussion

The seroprevalence of HEV varies depending on the geographic regions and to some extent to the sensitivity of serological assays, and characteristics of study population, including age. This study was conducted to investigate seroprevalence and viremia rates of HEV along with associated risk factors in high-risk groups in Turkey. The positivity rates for anti-HEV IgM, IgG, and HEV RNA were observed as 0%, 9.9%, and 0.6%, respectively. Two patients had viremia with HEV genotype 3, and both were initially seronegative for anti-HEV IgG and anti-HEV IgM. These findings revealed the importance and necessity of testing HEV RNA in patients suspected to have HEV infections in order to diagnose acute or chronic HEV infections. The independent risk factors for HEV seropositivity were eating undercooked meat and older age, regardless of the immune status of the patients.

In Europe, HEV3 is hyperendemic in southwest France and the rates of previous HEV exposure can reach up to 30% in Belgium, the Netherlands, and Germany (20). According to results of a systematic review, anti-HEV seroprevalence ranges from 0 to 12.4% among healthy population in Turkey. The prevalence was 7-8% in pregnant women, 13% in patients with chronic HBV infection, 54% in CHC patients, 13.9-20.6% in patients with chronic renal failure, and 35% among agriculture workers (21). HEV seroprevalence was reported as 33.4% among Turkish individuals immigrating form Turkey to the Netherlands and as similar to that in Dutch population (22). In a recently published multicentric study from Turkey, the HEV seroprevalence was investigated among blood donors (n=2011) using two different commercial ELISA assays. The anti-HEV seroprevalence was found 11.5% using Dia.Pro (Milan, Italy) assay and 12.2% using Wantai (Beijing, China) assay. HEV RNA was screened on anti-HEV antibody positive patients (n=272) and none of these patients was observed as viremic (23). For the first time in the literature, HEV epidemiology of RT recipients and allo-HSCT recipients in Turkey was investigated in this study by using both serology and PCR in all patients.

A recently published meta-analysis reported that anti-HEV seroprevalence in 14,626 transplant recipients varied between 6%-29.6% in different commercially available assays. Although there was a wide range for anti-HEV seroprevalence, HEV RNA positivity rate was 1.2% (95% CI: 0.9-1.6) among these patients (24). The rates of HEV RNA positivity were reported as 2.4% and 0.13% among patients having allo-HSCT or hematological malignancies in the Netherlands and the UK, respectively (25,26). Additionally, Ankcorn et al. (27) found that HEV RNA positivity was 0.66% among 2419 solid organ transplant recipients (1181 kidney, 869 liver, 229 heart, 110 lung, 21 kidney/liver, 6 heart/lung, 2 heart/kidney, and 1 lung/liver). In contrast, a large-scale study from the United Kingdom involving 225,000 blood donations demonstrated that 0.035% of recipients had HEV viremia and this rate was even lower (0.002%) in plasma donations in the US (28, 29). This difference between donors and immunocompromised patient groups can be explained by longer durations of HEV viremia caused by less effective HEV clearance in immunocompromised patients. In another study from Denmark, 4023 immunosuppressed patients were retrospectively tested for anti-HEV IgG and HEV RNA. HEV seroprevalence was 22.0% among all patients and increased with older age (65% increased risk per 10 years of increasing age). Only six patients (0.15%) were identified as HEV RNA positive and none of them did develop chronic HEV infection at follow-up. HEV RNA prevalence rates were 0.58% and 0.21% among allo-HSCT recipients and solid organ transplant recipients, respectively (30). To best of our knowledge, this is the first study investigating the seroprevalence of HEV among patients including RT recipients and allo-HSCT recipients in Turkey. The anti-HEV IgG positivity rate was 9.9% (29/292) in all patients with 3.8% (6/157) in RT group and 10.8% (5/46) in allo-HSCT group. These results appear to be parallel with the data of earlier studies and lower seroprevalence among RT recipients can largely be explained by immunosuppressive effects of drugs actively used to prevent organ rejection. All patients were tested for HEV RNA positivity, where only 2 patients (one RT recipient and one patient with AH) were identified as having HEV3 viremia. The rate of HEV RNA positivity was 0.6% both in whole patients and RT recipient subgroup. This rate was similar with that of the studies published from Europe.

The main transmission route for HEV-3 is supposedly foodborne, primarily from undercooked pig products. Additionally, environmental transmission routes from irrigation water or living in close proximity with farm animals have also been demonstrated as well-established risk factors for both healthy and immunosuppressed persons (31). In this study, eating undercooked meat was an independent risk factor to higher HEV seroprevalence. However, close contact with farm animals did not attain to statistical significance in multiple logistic regression analysis. This finding may be related to the lack of statistical power due to the small number of patients who have farm animal contact. Moreover, the farm animals in Turkey are mostly sheep and cows which may not have viral shedding as large as pigs. HEV3 infections have been assumed to be associated with the risk of development of chronic hepatitis in up to 50% of solid organ transplant recipients (32). However, in current study, it was not the case for the 2 patients with HEV3 viremia despite one being an RT recipient. As known, chronic HEV infections among solid organ transplant recipients are more likely to develop in deeply immunosuppressed patients. In particular, the CD2, CD3, and CD4 T cell subpopulations are significantly lower in patients with chronic HEV infections than in those who spontaneously clear HEV viremia (12). Unfortunately, data on T cell subpopulations and HEV RNA viral load were not available in our patient with RT. Other patient with HEV viremia was not immunocompromised and had acute HEV infection with accompanying biliary cholangitis.

In consistent with previous studies indicating that the seroprevalence was in close association with increasing age (33), we found an increased risk of anti-HEV seropositivity in older patients. Furthermore, age was an independent risk factor to anti-HEV IgG seropositivity in both multiple logistic regression analyses that were constructed for all patients and non-immunocompromised patients.

This study has strengths and limitations. It was designed as a cross-sectional study exploring not only HEV seropositivity rates but also HEV RNA viremia rates and risk factors for HEV exposure in high-risk individuals, including immunocompromised patients, in Turkey. For the first time in the literature, HEV genotype 3 was demonstrated in 2 patients from Turkey. Additionally, the prospective surveillance of these patients with HEV viremia had documented spontaneous clearance of HEV without development of chronic HEV infection at the visit undertaken 6 months after enrollment. As eating undercooked meat was a significant risk factor for seropositivity for HEV, the findings of this study reveal that possible foodborne HEV transmission routes should be investigated in Turkey. There were also some limitations of the study. First, this study was conducted in a single center. Therefore, the results of this study cannot be generalized over Turkish population. Secondly, data regarding the risk factors of HEV infection were collected by applying a face-to-face questionnaire. Hence, the risk for recall bias should be beard in mind while interpreting the results. In order to reduce the effect of the recall bias, the accuracy of data was also checked from the hospital database when possible. In addition, it cannot be ruled out that there may be additional confounding factors that were not included in this study. Last but not least, this is a point cross-sectional study in which serum samples were obtained at a random time during the course of RT and allo-HSCT. This may have led to an underestimation of both HEV viremia rates and chronic HEV infection.

In conclusion, HEV3 appears to be an emerging public health problem in Turkey. The results of this study strongly support the foodborne transmission of HEV as the principal transmission route regardless of immune status of patients. Since the patients with HEV viremia in this study were initially seronegative for anti-HEV IgG and anti-HEV IgM, we believe that testing HEV RNA is certainly required for diagnosing acute or chronic HEV infections in suspected patients.

## Data Availability

Data can be presented when required.

